# Prognostic and Immunological Significance of ARID1A Status in Endometriosis-Associated Ovarian Carcinoma

**DOI:** 10.1101/2021.09.16.21262993

**Authors:** Karolin Heinze, Tayyebeh M. Nazeran, Sandra Lee, Pauline Krämer, Evan S. Cairns, Derek S. Chiu, Samuel C.Y. Leung, Eun Young Kang, Nicola S. Meagher, Catherine J. Kennedy, Jessica Boros, Friedrich Kommoss, Hans-Walter Vollert, Florian Heitze, Andreas du Bois, Philipp Harter, Marcel Grube, Bernhard Kraemer, Annette Staebler, Felix K.F. Kommoss, Sabine Heublein, Hans-Peter Sinn, Naveena Singh, Angela Laslavic, Esther Elishaev, Alex Olawaiye, Kirsten Moysich, Francesmary Modugno, Raghwa Sharma, Alison H. Brand, Paul R. Harnett, Anna DeFazio, Renée T. Fortner, Jan Lubinski, Marcin Lener, Aleksandra Tołoczko-Grabarek, Cezary Cybulski, Helena Gronwald, Jacek Gronwald, Penny Coulson, Mona A El-Bahrawy, Michael E. Jones, Minouk J. Schoemaker, Anthony J. Swerdlow, Kylie L. Gorringe, Ian Campbell, Linda Cook, Simon A. Gayther, Michael E. Carney, Yurii B. Shvetsov, Brenda Y. Hernandez, Lynne R. Wilkens, Marc T. Goodman, Constantina Mateoiu, Anna Linder, Karin Sundfeldt, Linda E. Kelemen, Aleksandra Gentry-Maharaj, Martin Widschwendter, Usha Menon, Kelly L. Bolton, Jennifer Alsop, Mitul Shah, Mercedes Jimenez-Linan, Paul D.P. Pharoah, James D. Brenton, Kara L. Cushing-Haugen, Holly R. Harris, Jennifer A. Doherty, Blake Gilks, Prafull Ghatage, David G. Huntsman, Gregg S. Nelson, Anna V. Tinker, Cheng-Han Lee, Ellen L. Goode, Brad H. Nelson, Susan J. Ramus, Stefan Kommoss, Aline Talhouk, Martin Köbel, Michael S. Anglesio

## Abstract

ARID1A (BAF250a) is a component of the SWI/SNF chromatin modifying complex, plays an important tumor suppressor role, and is considered prognostic in several malignancies. However, in ovarian carcinomas there are contradictory reports on its relationship to outcome, immune response, and correlation with clinicopathological features. We assembled a series of 1623 endometriosis-associated ovarian carcinomas, including 1078 endometrioid (ENOC) and 545 clear cell (CCOC) ovarian carcinomas through combining resources of the Ovarian Tumor Tissue Analysis (OTTA) Consortium, the Canadian Ovarian Unified Experimental Resource (COEUR), local, and collaborative networks. Validated immunohistochemical surrogate assays for *ARID1A* mutations were applied to all samples. We investigated associations between ARID1A loss/mutation, clinical features, outcome, CD8+ tumor-infiltrating lymphocytes (CD8+ TIL), and DNA mismatch repair deficiency (MMRd). ARID1A loss was observed in 42% of CCOC and 25% of ENOC. We found no associations between ARID1A loss and outcomes, stage, age, or CD8+ TIL status in CCOC. Similarly, we found no association with outcome or stage in endometrioid cases. In ENOC, ARID1A loss was more prevalent in younger patients (p=0.012), and associated with MMRd (p<0.001), and presence of CD8+ TIL (p=0.008). Consistent with MMRd being causative of *ARID1A* mutations, in a subset of ENOC we also observed an association between *ARID1A* loss-of-function mutation as a result of small indels (p=0.011, vs. single nucleotide variants). In ENOC, the association between ARID1A loss, CD8+ TIL, and age, appears confounded by MMRd status. Although this observation does not explicitly rule out a role for ARID1A influence on CD8+ TIL infiltration in ENOC, given current knowledge regarding MMRd, it seems more likely that effects are dominated by the hypermutation phenotype. This large dataset with consistently applied biomarker assessment now provides a benchmark for the prevalence of *ARID1A* loss-of-function mutations in endometriosis-associated ovarian cancers and brings clarity to the prognostic significance.

## Introduction

Adenine-thymine rich interactive domain 1A (ARID1A) is a key member of the mammalian Switch/Sucrose Non-Fermentable (mSWI/SNF) chromatin remodeling complex which enables nucleosome conformation to promote DNA accessibility in an ATP-dependent process [1]. *ARID1A* encodes a protein which facilitates target-specific DNA binding. By binding to transcription activators or repressors, it regulates DNA transcription [2,3] and thus regulates a wide variety of cell cycle, differentiation, and development processes [4]. Recent experiments have also suggested that ARID1A interacts with regulators of the genomic stability/DNA damage and replication machinery [5–7]. *ARID1A* is the most frequently mutated subunit of the SWI/SNF complex detected in a broad spectrum of human cancers including gastrointestinal tract, lung, and breast malignancies [3,8,9]. Genome-wide sequencing analyses have identified *ARID1A* inactivation across ovarian malignancies, generally restricted to endometriosis associated ovarian carcinoma (EAOC) - clear cell (CCOC) and endometrioid ovarian carcinomas (ENOC). Loss-of-function (LOF) mutations in *ARID1A* have been reported in close to 50% of CCOC and 30% of ENOC [10–12]. In addition, genomic and functional studies have demonstrated that ARID1A inactivation promotes a malignant phenotype and is an early event in cancer pathogenesis; inactivation is broadly implicated in cancer progression, [13,14] yet it appears insufficient to drive transformation or tumor formation alone [10,15–20].

Somatic mutations in *ARID1A* are dominantly loss-of-function nonsense or frameshift mutations (insertion/deletion) that result in loss of protein expression and corresponding immunoreactivity in immunohistochemistry-based assessment. Therefore, the use of immunohistochemistry (IHC) to determine aberrant ARID1A protein expression and expression patterns is widely accepted and proven to work as a surrogate marker for *ARID1A* LOF mutations – as long as stringent methodology and appropriate antibody selection is adhered to [10,21]. While a consensus on IHC usage now exists, there are contradictory reports on ARID1A loss of protein expression/mutation with respect to outcomes (Table 1). Some groups have observed ARID1A depletion to be associated with favourable prognosis in a study of *ARID1A* mutation in the setting of impaired DNA (mismatch) repair mechanism in endometrial carcinomas. In contract, others suggest an adverse outcome relying on clinical prognostic features, and finally some studies fail to find associations in any way [22–26].

**Table 1.** Contradictory effects of ARID1A alterations reported in ovarian carcinoma literature.

|  |  |
| --- | --- |
| **ARID1A mutation is associated with tumor promoting properties** |  |
| ARID1A mut correlates with poor prognosis, lower survival, high degree of mutability, chemotherapy resistance and early reoccurrence, progression, earlier disease onset | [23,25,26,35,52,76-80] |
| **ARID1A mutation has no effect** |  |
| ARID1A mut shows no survival difference, no difference in stages, no correlation to chemo sensitivity | [17,24,40,81-83] |
| **ARID1A mutation is associated with tumor suppressive properties** |  |
| ARID1A mut enhances patient survival, differentiation of epithelia, lack of mut results in unfavorable outcome | [53-55] |

The correlation of *ARID1A* mutational status with outcomes in different cancer types is varied. ARID1A inactivation correlates with poor outcome in breast, bladder, and bone malignancies [9,27,28] while it has no apparent prognostic significance in esophageal adenocarcinoma [29]. In endometrial carcinoma *ARID1A* LOF mutations are seen as favorable prognostic biomarker, though potentially confounded with impaired DNA mismatch repair (MMR) [30,31], and a target for development of new therapeutic interventions [32–34].

In ovarian carcinomas the prognostic significance of *ARID1A* LOF mutation remains equivocal. Conflicting reports have implied positive, negative, and lack of any association with outcome and clinical features [25–29]. ARID1A deficiency has been linked to elevated tumor infiltrating lymphocytes (TIL) within specific histotypes, however sample sizes in these studies are relatively small [35–37]. Confirmation of such a finding may be important for clinical management as TIL are associated with improved outcomes in many solid tumors including ovarian carcinomas, though the strongest effects are observed in HGSOC. Beneficial CD8+ TIL effects are notable in ENOC, where ARID1A alterations are also prevalent, though they do not appear to be as strong as trends observed in HGSOC [38]. Likewise, prognostic CD8+ TIL effects in other histotypes have not been observed though small sample sizes have hindered efforts. Poor response to conventional treatment in advanced EAOC and emerging evidence of benefits of immunomodulatory therapies in EAOC treatment suggests there may be considerable value in validating biomarker predictive of their activity [39,40]. As noted above, ARID1A alterations appear enriched in DNA mismatch repair deficient (MMRd) endometrial carcinomas, a feature also common in ENOC, and generally associated with improved outcomes [22,30,41–43].

Herein, we leverage collective and consortia-based ovarian carcinoma tissue collections from the Ovarian Tumour Tissue Analysis Consortium, the Canadian Ovarian Experimental Unified Resource, local resources, and collaborations to investigate the prognostic and clinicopathological association of ARID1A loss-of-function alteration in ovarian carcinomas. We also re-visit CD8+ TIL infiltration and MMRd status, published previously [38,43], now in the context of ARID1A LOF ovarian carcinoma.

## Materials and Methods

### Sample collection

A total of 5115 ovarian carcinoma cases of all histotypes were initially included with detailed examination on a subset of 1623 EAOC including 1078 ENOC and 545 CCOC. Participating studies included collections from the Ovarian Tumor Tissue Analysis (OTTA) consortium (CCOC=386, ENOC=558; and all non-EAOC cases n=2598), the Canadian Ovarian Experimental Unified Resource (COEUR) repository (ENOC=156), local Vancouver cohort (VAN) (ENOC=217, CCOC=159) and from German collaborators (Tübingen, Essen, Heidelberg, Friedrichshafen; ENOC=147). The project was conducted in compliance with the Canadian Tri-Council Policy Statement on Ethical Conduct for Research Involving Humans (TCPS2, 2018); usage of specimen and associated clinical data was approved by local institutional research ethics boards (see also Table S1). Histotype review and confirmation on the OTTA and COEUR cohorts has been described previously, including both central and histotype-specific IHC [44]. All other samples were pathologist reviewed and subject to confirmation using histotype specific IHC [43,45]. Detailed information regarding sample collections, tissue microarray (TMA) construction, pathological variables, and clinical data of participants in the OTTA, COEUR and VAN and German cohort have been described previously [43,46,47].

### Immunohistochemistry and Scoring

IHC was performed on TMA for ARIDA1 (details in Supplemental methods). A subset of cases from COEUR, VAN and Germany were stained for MMR proteins (MLH1, PMS2, MSH2, MSH6) (Figure S1). The immunohistochemical method and scoring has been provided in prior published studies [43,48] (see also Supplemental methods). CD8+ TIL scores for OTTA samples were taken from Goode *et al*. [38] wherein staining and scoring was replicated in the same lab and pathologist (MK) for additional cohorts. Regarding the interpretation, ARID1A was assessed for nuclear staining in tumour epithelium (absent, present, subclonal) with retained stromal nuclear staining serving as an obligate internal control. During analysis ARID1A absence and subclonal scores were merged (unless otherwise noted in the text). Samples were considered MMRd if loss of staining in any of the four MMR markers was observed (with retained stromal staining serving as an obligate internal control [43]). CD8+ TIL infiltration was binned by counting and averaging the number of CD8+ cells in the tumor epithelium of TMA cores (0=none, low=1-2, moderate=3-19, high ≥20 [38]).

### Statistical analysis

Statistical analyses were conducted separately for ENOC and CCOC. Grade 1 was considered low and grades 2-4 as high grade. For stage-stratified analysis FIGO I and II were defined as low-stage and FIGO III and IV as high-stage. Welch’s one-way or Chi Square tests were used to evaluate univariable association for continuous and categorical data, respectively. Clinical follow up was left truncated and, if exceeding 10 years, right censored as at December 31st of the 10th year post diagnosis to minimize ascertainment bias and ensure non-informative censoring. The survival outcome measures, overall, disease-specific, and progression-free survival were assessed via Kaplan-Meier plotting and statistical significance was determined using a log-rank test. To calculate the multivariable effect of ARID1A LOF and other clinicopathological parameters on the survival outcome, Cox proportional hazards models were applied. In situations where more than 80% censoring occurred within a minimum of one variable, hazard estimates were generated using the Firth bias reducing correction. *P* values of < 0.05 were considered significant.

## Results

### ARID1A loss is a feature of endometriosis-associated ovarian histotypes

A total of 5115 cases were examined by *ARID1A* mutation surrogate IHC across all major histotypes (Table S2). Clinicopathological variables including age, stage, grade (ENOC only) and residual disease were factored into overall (OS), progression-free (PFS) and disease-specific (DSS) survival. Our combined cohort is not population based and is deliberately enriched for ENOC specific sub-analysis discussed below: 2% of cases were classified as low (LGSOC), 51% high-grade serous ovarian carcinoma (HGSOC), 21% as ENOC, 11% as clear cell ovarian carcinoma (CCOC), 5% as mucinous ovarian carcinoma (MOC) and 10% as other (including borderline tumors) (Table S2). Protein expression for ARID1A was detected in ≥94% of LGSOC, HGSOC, and MOC, while ARID1A loss was most prevalent in ENOC (25%; including subclonality in 19 cases, or 2% of all) and CCOC (42%; including subclonality in 7 cases, or 1% of all). ARID1A loss was significantly enriched in EAOC (p<0.001) (Figure 1A).

**Figure 1.**
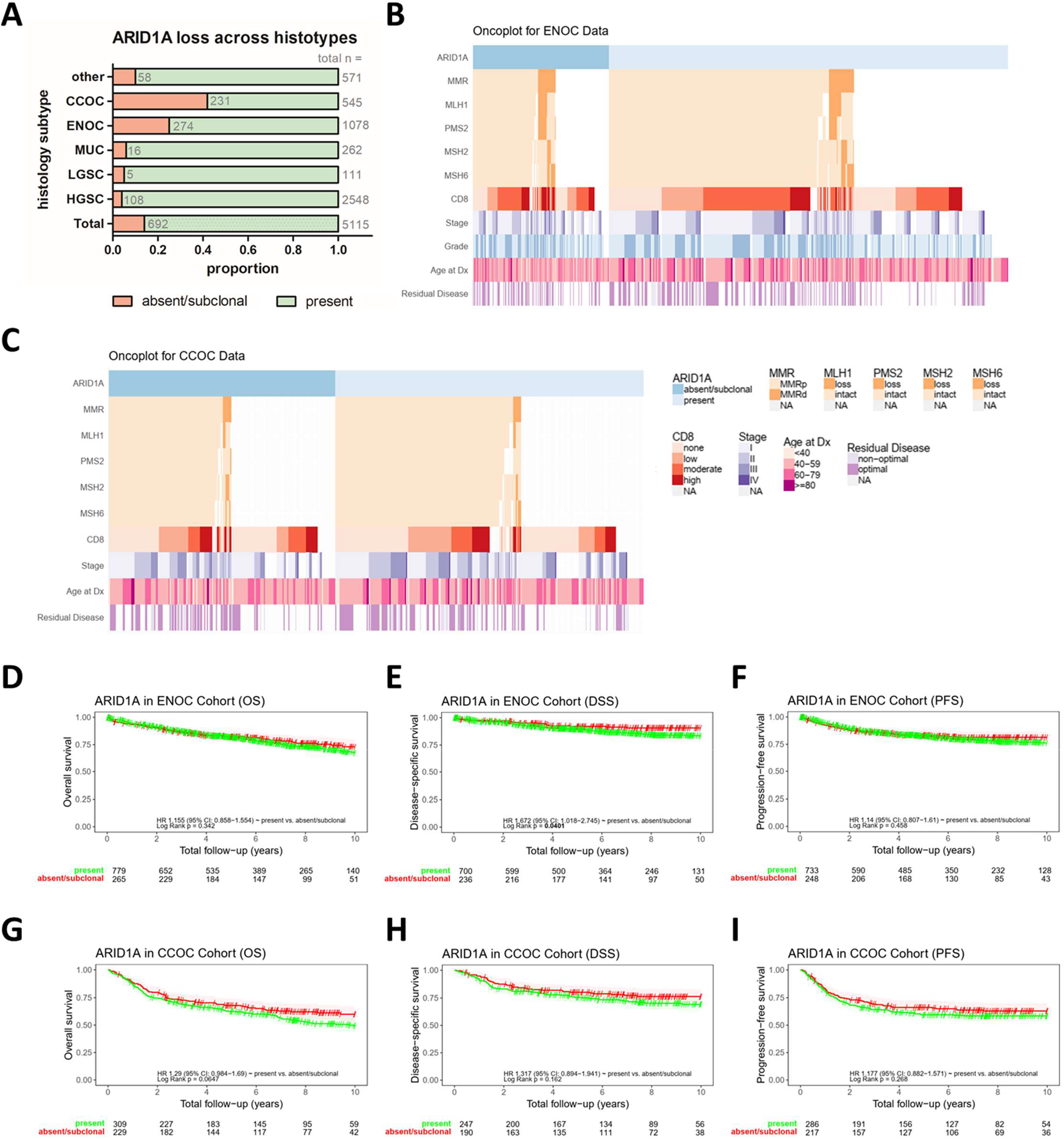
Results from ARID1A classification in ovarian carcinomas. **A** Frequency of ARID1A loss detected in major ovarian carcinoma histological subtypes; ‘other’ contain cases of mixed cell, borderline and other specified epithelial ovarian tumors, **B**,**C** Oncoplot outlining full ENOC (**B**) and CCOC (**C**) cohort of cases (in columns) along with ARID1A, MMR and CD8+ status and clinicopathologic features, **D-F** Kaplan-Meier survival curves depicting OS (n=1044), DSS (n=936) and PFS (n=575) with absent vs. present ARID1A status in ENOC, **G-I** Kaplan-Meier survival curves depicting OS (n=538), DSS (n=437) and PFS (n=322) with absent vs. present ARID1A status in CCOC.

### Clinicopathological associations within EAOC cohorts

Our sample set appeared consistent with expected clinicopathological associations across stage, grade (considered for ENOC only), and residual disease in ENOC and CCOC in univariable analysis (Figure 1B-C; Table 2; Figure S2, S3). In multivariable survival analysis stage and grade were significantly associated with all outcome endpoints in ENOC (OS, DSS, and PFS: all p<0.05, Table 3A). Grade retained significance when the analysis was limited to low-stage (FIGO I/II) ENOC (OS, DSS, PFS all p<0.05, Table 3B). Presence of residual disease was associated with reduced DSS (p=0.026) and PFS (p=0.011) which again was consistent when examining PFS in only high-stage (FIGO III/IV) (p=0.002, Table 3C). Across CCOC, in multivariable survival analysis stage and residual disease were significantly associated with outcome endpoints when examining all cases or somewhat when restricting to low-stage (stage all p<0.001; residual disease all OS and PFS p<0.05, low-stage OS p=0.034, Table 3A+B). Age was independently associated with all survival outcomes in CCOC regardless of looking at the full cohort (p<0.01, Table 3A), restricting to high-stage (p<0.05) (Table 3C), or OS in low-stage (p=0.038, Table 3B).

**Table 2.** Univariable Survival in EAOC. Summary of p-Values obtained during univariable survival analysis in ENOC and CCOC on clinicopathological and molecular parameters. Corresponding Kaplan-Meier curves are indicated and can be found in figure 1 and supplementary figures 2 and 3.

| **ENOC** | **OS** | **DSS** | **PFS** | **cf. (log-rank)** |
| --- | --- | --- | --- | --- |
| age | **0.001** | **<0.001** | **<0.001** |  |
| ARID1A | 0.892 | 0.135 | 0.525 | Fig.1 D-F |
| Grade | **<0.001** | **<0.001** | **<0.001** | Fig.S2 A, E, I |
| Stage | **<0.001** | **<0.001** | **<0.001** | Fig.S2 B, F, J |
| Residual disease | **<0.001** | **<0.001** | **<0.001** | Fig.S2 D, H, K |
| MMR status | 0.237 | 0.974 | 0.965 | Fig. S4 A-C |
| CD8+ status | **0.034** | 0.198 | 0.086 | Fig.S3 A-C |
| **CCOC** |  |  |  |  |
| age | **<0.001** | **<0.001** | **<0.001** |  |
| ARID1A | 0.297 | 0.316 | 0.392 | Fig.1G-I |
| Stage | **<0.001** | **<0.001** | **<0.001** | Fig.S3 A, D, G |
| Residual disease | **<0.001** | **0.001** | **<0.001** | Fig.S3 C, F, I |
| MMR status | 0.868 | 0.505 | 0.459 |  |
| CD8+ status | 0.377 | 0.522 | 0.324 | Fig.S3 D-F |

**Table 3.** Multivariable Survival in EAOC. **A** Multivariable survival model statistics when considering clinicopathological parameters and ARID1A for OS, DSS and PFS in the full ENOC and CCOC cohorts, (**B – C)** Multivariable survival model statistics in only low-(**B**) and only high-staged (**C**) ENOC and CCOC cohorts respectively.

| **A** |  |  | **OS** |  | **DSS** |  | **PFS** |  |
| --- | --- | --- | --- | --- | --- | --- | --- | --- |
|  |  |  | HR<br>(95%CI) | LRT<br>PValue | HR<br>(95%CI) | LRT<br>PValue | HR<br>(95%CI) | LRT<br>PValue |
| **ENOC** | *# events/n* |  | 78/368 |  | 37/334 |  | 76/360 |  |
| Age |  |  | 1.05 | 0.299 | 1.02 | 0.835 | 1.25 | <0.001 |
| Stage | II |  | 1.53 |  | 1.34 |  | 2.84 |  |
|  | III |  | 3.71 | <0.001 | 3.17 | 0.017 | 4.14 | <0.001 |
|  | IV |  | 8.8 |  | 8.59 |  | 12.78 |  |
| Grade | high |  | 2.14 | 0.003 | 4.05 | <0.001 | 2.41 | <0.001 |
| residual | optimal |  | 0.56 | 0.056 | 0.38 | 0.026 | 0.47 | 0.011 |
| ARID1A | present |  | 1.03 | 0.929 | 1.04 | 0.931 | 1.13 | 0.669 |
| **CCOC** | *# events/n* |  | 106/211 |  | 40/152 |  | 96/184 |  |
| Age |  |  | 1.15 | 0.001 | 1.22 | 0.006 | 1.52 | <0.001 |
| Stage | II |  | 1.48 |  | 1.06 |  | 1.9 |  |
|  | III |  | 4.88 | <0.001 | 6.19 | <0.001 | 7.5 | <0.001 |
|  | IV |  | 9.56 |  | 13.87 |  | 26.93 |  |
| residual | optimal |  | 0.55 | 0.030 | 0.7 | 0.444 | 0.43 | 0.005 |
| ARID1A | present |  | 0.75 | 0.185 | 0.6 | 0.162 | 0.94 | 0.783 |

| **B** |  |  |  |  |  |  |  |  |
| --- | --- | --- | --- | --- | --- | --- | --- | --- |
| **Low Stages EAOC (FIGO I + II)** |  |  |  |  |  |  |  |  |
| **LowENOC** | *# events/n* |  | 45/302 |  | 19/282 |  | 42/296 |  |
| Age |  |  | 1.04 | 0.528 | 0.98 | 0.859 | 1.28 | 0.001 |
| Stage | II |  | 1.58 | 0.190 | 1.22 | 0.720 | 2.96 | 0.002 |
| Grade | high |  | 2.29 | 0.012 | 3.41 | 0.013 | 2.19 | 0.016 |
| residual | optimal |  | 0.69 | 0.496 | 0.28 | 0.085 | 0.68 | 0.465 |
| ARID1A | present |  | 1.12 | 0.763 | 0.94 | 0.917 | 1.07 | 0.842 |
| **lowCCOC** | *# events/n* |  | 53/148 |  | 18/118 |  | 44/139 |  |
| Age |  |  | 1.12 | 0.038 | 1.16 | 0.115 | 1.61 | <0.001 |
| Stage | II |  | 1.34 | 0.314 | 1.17 | 0.762 | 1.79 | 0.080 |
| residual | optimal |  | 0.36 | 0.034 | 7.7e+07 | 0.161 | 0.35 | 0.069 |
| ARID1A | present |  | 0.84 | 0.566 | 0.69 | 0.457 | 0.77 | 0.424 |

| **C** |  |  |  |  |  |  |  |  |
| --- | --- | --- | --- | --- | --- | --- | --- | --- |
| **High Stage EAOC (FIGO III + IV)** |  |  |  |  |  |  |  |  |
| **highENOC** | *# events/n* |  | 33/66 |  | 18/52 |  | 34/64 |  |
| Age |  |  | 1.06 | 0.500 | 1.03 | 0.809 | 1.31 | 0.011 |
| Stage | IV |  | 2.18 | 0.211 | 1.32 | 0.751 | 1.49 | 0.596 |
| Grade | high |  | 1.59 | 0.280 | 4.77 | 0.041 | 2.54 | 0.047 |
| residual | optimal |  | 0.47 | 0.085 | 0.38 | 0.133 | 0.24 | 0.002 |
| ARID1A | present |  | 0.87 | 0.819 | 1.31 | 0.782 | 0.83 | 0.783 |
| **highCCOC** | *# events/n* |  | 53/63 |  | 22/34 |  | 53/59 |  |
| Age |  |  | 1.31 | 0.002 | 1.55 | 0.002 | 1.46 | 0.022 |
| Stage | IV |  | 2.14 | 0.270 | 3.52 | 0.244 | 4.7e09 | 0.006 |
| residual | optimal |  | 0.69 | 0.267 | 0.43 | 0.196 | 0.32 | 0.010 |
| ARID1A | present |  | 0.66 | 0.266 | 0.86 | 0.802 | 1.37 | 0.504 |

### Clinicopathological associations of ARID1A loss within EAOC cohorts

In univariable non-parametric analysis, no association between ARID1A loss and the clinicopathological variables stage, grade (ENOC), or residual disease was seen in ENOC or CCOC (Table S4A, S4B), and this was consistent with respect to grade and residual disease when considering low-stage and high-stage cases separately. In low-stage ENOC, FIGO I was less commonly associated with ARID1A loss than in FIGO II cases (24% vs. 31%, p=0.034, data not shown) Amongst ENOC, ARID1A loss was significantly associated with a younger age of diagnosis, regardless of stage (mean age 55 vs. 57 respectively; p=0.012, Table S4A). ARID1A loss was associated with younger age at diagnosis in high-stage CCOC cases (mean 52 vs. 58; p=0.001, data not shown), but not low-stage CCOC. ARID1A loss status showed no significant associations with OS or PFS (p>0.05) in ENOC (Figure 1D-F). A significant association was observed for DSS in ENOC in univariable analysis (p=0.035). Of note, the subset of cases with DSS reported (n=936/1078; 87% of the cohort) showed a trend to lower stage and proportion of cases with optimal debulking compared to the complete case set analyzed for OS. In CCOC, we observed no association of OS, DSS, or PFS with ARID1A (p>0.1) (Figure 1G-I). No correlation between ARID1A loss and survival in either EAOC histotypes was observed in stage-based sub-analyses (Table S3). ARID1A loss was not an independent prognostic factor upon multivariable analysis for either ENOC or CCOC (Table 3).

### Association of ARID1A loss with CD8 TIL in CCOC and ENOC

We re-examined published data for the OTTA cohort [38] and newly generated CD8 IHC data on all other cases performed with the same assays and interpretation. CD8+ TIL scores were available for 933 ENOCs and 480 CCOCs and amongst those 26% of ENOCs and 44% of CCOCs showed an ARID1A loss (Figure 1B-C). In ENOC the loss of ARID1A was significantly associated with the presence of CD8+ TIL (p=0.008). The proportion of tumors with ARID1A loss increased with increasing levels of CD8+ TIL amongst ENOC: from 22% loss in CD8+ TIL negative to 37% loss in CD8+ TIL ≥20 (Table 4A; p=0.008). Notably, CD8+ TIL was also associated with MMR status amongst ENOC (p<0.001; Table 4A; see also below). In CCOC we also observed a slight increase in the proportion of cases with ARID1A loss as CD8+ TIL increased; however, this association did not reach statistical significance (Table 4B). Similarly, while we did observe a significant trend of increasing CD8+ TIL with MMRd in CCOC, it should be noted that there were few observed cases with MMRd in CCOC (5% overall). We did not observe any association between CD8+ TIL and grade (for ENOC) or stage in either histotype (Table 4). Kaplan-Meier and univariable survival analysis failed to show any significant influence of CD8+ TIL on DSS and PFS in either ENOC or CCOC outcomes (Figure S4A, Table S5). However, amongst ENOC, CD8+ TIL high (≥3) patients showed a modest, yet significant, improvement in OS compared to patients with lower levels (≤2) of CD8+ TIL (p=0.002). No trend in OS was seen in CCOC. In stage stratified sub-analysis CD8+ TIL was associated with OS and PFS in low-stage ENOC (p<0.05) and with OS, DSS, and PFS in high-stage CCOC (p<0.05, data not shown).

**Table 4.** Influence of CD8+ in EAOC. Univariable association between CD8+ status and clinicopathological parameter and ARID1A status in ENOC (**A**, n= 933) and CCOC (**B**, n=480).

| **A** | Variable | Levels | none | low | moderate | high | Total | PValue |
| --- | --- | --- | --- | --- | --- | --- | --- | --- |
|  | **Total** | N (%) | 264 (24%) | 189 (18%) | 351 (33%) | 129 (12%) | 1078 (100%) |  |
|  | **ARID1A** | absent/subclonal | 57 (22%) | 48 (25%) | 85 (24%) | 48 (37%) | 238 (26%) | **0.008** |
|  |  | present | 207 (78%) | 141 (75%) | 266 (76%) | 81 (63%) | 695 (74%) |  |
|  | **MMR** | MMRp | 147 (93%) | 110 (85%) | 233 (90%) | 62 (69%) | 552 (87%) | **<0.001** |
|  |  | MMRd | 11 (7%) | 19 (15%) | 27 (10%) | 28 (31%) | 85 (13%) |  |
|  |  | Missing | 106 | 60 | 91 | 39 | 296 |  |
|  | **Grade** | low grade | 142 (59%) | 95 (56%) | 201 (60%) | 66 (57%) | 504 (59%) | **0.828** |
|  |  | high grade | 98 (41%) | 75 (44%) | 134 (40%) | 49 (43%) | 356 (41%) |  |
|  |  | Missing | 24 | 19 | 16 | 14 | 73 |  |
|  | **Stage** | I | 118 (54%) | 98 (59%) | 179 (58%) | 65 (59%) | 460 (57%) | **0.643** |
|  |  | II | 60 (27%) | 40 (24%) | 90 (29%) | 30 (27%) | 220 (27%) |  |
|  |  | III | 38 (17%) | 23 (14%) | 32 (10%) | 13 (12%) | 106 (13%) |  |
|  |  | IV | 4 (2%) | 4 (2%) | 6 (2%) | 3 (3%) | 17 (2%) |  |
|  |  | Missing | 44 | 24 | 44 | 18 | 130 |  |

| **B** | Variable | Levels | none | low | moderate | high | Total | PValue |
| --- | --- | --- | --- | --- | --- | --- | --- | --- |
|  | **Total** | N (%) | 243 (45%) | 108 (20%) | 66 (12%) | 63 (12%) | 545 (100%) |  |
|  | **ARID1A** | absent/subclonal | 103 (42%) | 45 (42%) | 31 (47%) | 30 (48%) | 209 (44%) | **0.795** |
|  |  | present | 140 (58%) | 63 (58%) | 35 (53%) | 33 (52%) | 271 (56%) |  |
|  | **MMR** | MMRp | 136 (99%) | 78 (98%) | 34 (92%) | 32 (80%) | 280 (95%) | **<0.001** |
|  |  | MMRd | 2 (1%) | 2 (2%) | 3 (8%) | 8 (20%) | 15 (5%) |  |
|  |  | Missing | 105 | 28 | 29 | 23 | 185 |  |
|  | **Stage** | I | 109 (53%) | 46 (47%) | 23 (43%) | 24 (43%) | 202 (49%) | **0.822** |
|  |  | II | 56 (27%) | 26 (27%) | 17 (31%) | 20 (36%) | 119 (29%) |  |
|  |  | III | 39 (19%) | 25 (26%) | 13 (24%) | 11 (20%) | 88 (21%) |  |
|  |  | IV | 2 (1%) | 1 (1%) | 1 (2%) | 1 (2%) | 5 (1%) |  |
|  |  | Missing | 37 | 10 | 12 | 7 | 66 |  |

### ARID1A loss correlation with DNA mismatch repair status in ENOC

MMR status was available in 661 of 1078 ENOC cases. Absence of IHC staining in one or more of the evaluated MMR proteins (MLH1, MSH2, MSH6, PMS2) was seen in 87 cases and 36 exhibited both MMRd and ARID1A loss. 22% of all ENOC-ARID1A loss cases had concurrent MMR deficiency resulting in a significant association between ARID1A and MMR status (p<0.001; Figure 1B, Table S6).

Evaluating the specific MMR protein patterns, an enrichment between ARID1A loss and MLH1 and PMS2 loss was observed, even when only considering low-stage ENOC (p=0.013 and p<0.001, respectively, Table S6). This significant association is characterised by a 2.2 to 4-fold increased likelihood of detecting loss of MLH1 or PMS2 in combination with ARID1A loss compared to those that retained ARID1A staining. MMR status did not influence any measured outcome parameters (OS, DSS, PFS p>0.5, Table S7, Figure S5). Examination of *ARID1A* genetic variants, where data was available from previous studies [10,49,50], also showed *ARID1A* indel mutation were more commonly the cause of LOF mutations in *ARID1A* amongst MMRd ENOC compared to MMRp ENOC (p=0.011, Table S8).

Given the association of ARID1A loss with MMRd status we further examined this MMRd ENOC subset (n=87). Within the MMRd subset ENOC cases affected by ARID1A loss tended to be younger than MMRd ENOC with retained ARID1A expression, though not statistically significant (mean 52 vs. 56 y/o, p=0.129, Table S9A). No age difference was observed amongst MMR-proficient (MMRp) ENOC (mean 56 vs. 57 y/o; p=0.251, data not shown). There was no correlation with tumor grade or stage (p>0.5, Table S9A) albeit the sample size was small. As *ARID1A* indel mutations were more prevalent in MMRd ENOC (Table S8) we also considered that, if MMRd was causative of *ARID1A* mutations, one might expect a higher frequency of subclonal ARID1A alterations in this group. However, with only 19 cases showing subclonal ARID1A staining amongst ENOC our dataset was likely insufficient and only 3 were MMRd; no significant association was observed.

We also examined the relationship of clinicopathological features and survival within MMRd ENOC. Residual disease was of borderline significance for DSS (p=0.041, but not OS or PFS; Table S9B). Stage had a greater influence and was significant (p≤0.01) for OS, and PFS (Table S9B). We were not able to discern differences in outcome related to other variables (age, grade, ARID1A, or CD8+ TIL) within this comparably small subset of MMRd ENOC (Table S9B). In the larger subset of MMRp ENOC nearly all variables reflected trends seen with the full cohort and significant associations were observed for OS, DSS, and PFS across grade, stage, and residual disease. In MMRp ENOC age retained significance for DSS and PFS, and DSS only for CD8+ TIL status (p<0.05; Table S10).

## Discussion

In the present study, the prognostic significance of ARID1A loss-of-function was evaluated in a cohort of 1623 endometriosis-associated ovarian carcinoma patients with clinicopathological variables, MMR status and CD8+ TIL included as additional biomarkers of interest. Our series appears to represent the largest data set reporting on prognostic significance of ARID1A in (endometriosis-associated) ovarian carcinomas to date and is important to clear confusion on clinicopathological and prognostic associations of ARID1A loss, as a surrogate for LOF mutations.

As expected, clinicopathological features such as stage, grade (only for ENOC), and residual disease were confirmed as prognostic factors in EAOC. Our results are consistent with published data on the frequency of ARID1A alterations but now provided a more accurate benchmark: CCOC (42%) and ENOC (25%) [10–12]. Differences in *ARID1A* mutation frequencies relative to other reports may be due to smaller cohort sizes in other reports. Alternatively, because IHC reflects loss-of-function mutations, it is potentially unable to capture other non-synonymous somatic substitution mutations with unknown pathological effects and relevance. Nonetheless our data are in line with other emerging large-scale sequencing-based reports on CCOC (manuscript in prep [51]).

Our report confirms that ARID1A loss, as a surrogate for mutation status, has no impact on OS, or PFS in ENOC or CCOC. This is in contrast with other smaller scale studies suggesting either positive [23,37,52] or negative prognostic relationship for ARID1A loss in EAOCs [53–55]. It is also in contrast to findings in other cancer types where ARID1A inactivation has been depicted as a predictor of poor prognosis [56,57]. At least some of these prior studies in other cancers are small or moderate in size, with higher potential for type 1 errors, thus suggesting that larger studies with cancer-type context may be warranted [58].

Previous reports have suggested ARID1A loss is more prevalent in younger CCOC [25]. We partially validated this finding in our larger cohort of CCOC; however, ARID1A loss was only associated with younger age amongst high-stage CCOC. We also found ARID1A loss was more prevalent in younger aged ENOC overall. However, the younger age at diagnosis amongst ENOC patients was confounded by MMR status; it is already known that MMRd status is associated with younger age at diagnosis [43]. Herein, acquisition of an *ARID1A* loss-of-function mutation in the context of MMRd ENOC may be an effect modifier, and we observed that such cases retain an association with younger age at diagnosis compared to MMRd ENOC without ARID1A loss. ARID1A loss has been hypothesized to increase cell proliferation and likewise restoration of wildtype ARID1A appears to be more proliferation rate-limiting [59,60]. One may speculate that MMRd/ARID1A loss lesions may grow more rapidly, yet remain restricted to an ovarian microenvironment [61], resulting in more prevalent symptoms and younger discovery. This hypothesis is clearly dependent on very specific features of differentiation and somatic molecular background. Context-specific functional studies will be needed to deconvolute ARID1A functions in these cases. Earlier ENOC onset is typical for Lynch syndrome carriers whose disease is characterized by dysfunction in MMR pathway and is readily detectable in gynecological malignancies [62]. The influence of Lynch carriers on the younger age of MMRd ENOC may not be the sole explanation for this group’s younger age at diagnosis. Loss affecting MSH2, MSH6, or PMS2 (in the presence of intact MLH1) affected 49 out of 104 MMRd EAOC (39 ENOC, 10 CCOC) cases of probable Lynch syndrome. However, germline testing was not carried out in our cohort, and a recent analysis of germline MMR defects confirmed Lynch syndrome in 8/12 probable cases (67%) [63,64], none amongst those with concurrent PMS2/MLH1 loss by IHC.

We report a strong association between ARID1A loss and MMR deficiency, consistent with prior studies, including those investigating endometrial and ovarian endometrioid carcinomas [12,13,65–67]. Likewise, our data suggests MMRd may precede, and be causative of indel type *ARID1A* LOF mutations in MMRd ENOC. Despite this connection we did not observe an association between subclonal staining of ARID1A and MMRd, as might be expected. However, our TMA-based approach is not well suited to detect subclonal staining patterns and we lacked deep sequencing data on the MMRd subset of cases that would be definitive of subclonal *ARID1A* alterations.

The association of ARID1A loss and CD8+ TIL infiltration in ENOC was also confounded by MMR status. We found that ARID1A loss in ENOC did correlate with increasing CD8+ TIL density, and while TIL density was prognostic [38], ARID1A status was not. The association between MMRd and increasing CD8+ TIL density was also highly significant. As high neoantigen load generated by the hyper-mutation MMRd phenotype is well recognized to drive immune infiltration, this may be the more dominant driver for this observation [50]. Furthermore, MMRd is relative uncommon in CCOC with 5% in our cohort and 1.7% in a recent series [64].

Two recent studies suggested that ARID1A depleted ovarian tumors had increased CD8+ TIL infiltration [35,37]. It should be noted both studies’ scoring parameters for CD8 or ARID1A were not presented, their ARID1A IHC did not use a validated antibody known to act as a surrogate of LOF mutations [21], and one of the study cohorts was a mix of CCOC and HGSOC [35]. Our standardized approach, and large-scale analysis, did not result in any significant relationship between ARID1A loss and CD8+ TIL infiltration amongst CCOC. In fact, high CD8+ TIL infiltration amongst CCOC was observed in only 12% of our cohort [38]. This is a striking difference to the proportion of ARID1A “low” and CD8 “high” which may have been as much as 74% in the Shen *et al*. report or the 20-30% CD8 “positive” reported for *ARID1A* mutant/loss in the Kuroda report [35,37]. Both Shen *et al*. and Kuroda *et al*. suggest *ARID1A* mutations are associated with higher tumor mutation burden in CCOC, with Shen *et al*. suggesting this is due to lost interactions with the MMR complex and subsequent phenotypic deficiency in MMR. Our current study did not examine tumor mutation burden, though prior work may support a slight increase with *ARID1A* mutation [50]; however, the mutation signatures in CCOC are not consistent with a pattern that would be expected (i.e. COSMIC signatures 6, 15, 20 and/or 26) if *ARID1A* loss leads to deficiencies in DNA mis-match repair or a hypermutation phenotype [50].

As endometriosis-associated ovarian cancers account for only ∽ 25% of ovarian carcinomas, studying these rarer ovarian histotypes can be particularly challenging. A strength of the present study was the availability of a large number of EAOC cases (n=1623) through local and collaborative/consortia-based collections that have undergone extensive review and immunohistochemical validation of histology. 545 CCOC and 1078 ENOC cases were stained for ARID1A LOF and sub-analyses of MMR and CD8+ TIL status were performed. We also applied centralized and uniform immunohistochemical assays with validated scoring parameters. In particular, we have benefitted from the sequence-based validation of ARID1A IHC as a mutation surrogate and a well-developed standard for characterization of MMR status [10,21,49]. Earlier study findings may have been affected by a lack of consensus on applied scoring systems and mixed usage of varying commercially available antibodies, as noted above.

We recognize that our study is limited by the use of TMA based resources with reduced ability to detect subclonal staining for our biomarkers of interest. In fact, varied expression patterns of ARID1A within a tumor might influence the prognosis in gastric cancer [68] and these subclonal patterns may result in clonal expansion and progressive tumor evolution [12]. We also recognize that CD8+ TIL infiltration was captured only in a semi-quantitative fashion, though given availability of historical data we felt it was justifiable to maintain the previous scoring system for comparability and our data appears to be in line with more quantitative reports [69–71]. CD8+ TIL infiltration is also a relatively limited picture of immune infiltration. While CD8+ TIL have shown to be prognostic in HGSOC it is entirely reasonable to assume other immune populations are more important to the establishment, and progression of EAOC. Such a hypothesis is consistent with the recent report of a (prognostic) subset of CCOC with a high-immune/inflammatory gene expression profile (so called “MesCC”), a profile that the authors point out is not driven by CD8+ TIL-specific signatures [36]. Additionally, we are aware that despite the large cohort size overall, subset analysis of MMRd ENOC is underpowered making conclusive statements difficult within this group. Further expansion of our cohort and collection of additional molecular data would be favourable, especially for ENOC where inclusion of *POLE* mutation status was not possible due to the nature of available samples (in TMA format only). Including *POLE* status would have enabled the entire cohort to be examined in the context of modern molecular subtypes of endometrioid carcinomas [43]. Likewise, such subtype classification structure has not been widely explored in CCOC and more rigorous molecular analysis including mutational biomarkers and/or epigenetic states may be highly valuable in future analyses of this cohort. Finally, it should be noted that the lack of prognostic significance for ARID1A does not discount continued development of direct targeting, synthetic lethal strategies, and/or investigation into predictive value of ARID1A for immune-modulatory therapies [72–75]. As one of the most prevalent biomarkers in EAOCs it is a potentially high value biomarker with mechanisms of action that may be dependent on molecular context of the cancer (and subtype) in which alterations occur. Future, well-warranted, theragnostic development around ARID1A must take note of potential confounders and molecular context.

## Supporting information

Table S1 (cohort details)

## Data Availability

All data is contained within the manuscript and or associated supplemental information.

## Abbreviations

CCOC: clear cell ovarian carcinoma
COEUR: Canadian Ovarian Unified Experimental Resource
EAOC: endometriosis-associated ovarian carcinoma
ENOC: endometrioid ovarian carcinoma
HGSOC: high grade serous ovarian carcinoma
IHC: immunohistochemistry
LOF: loss-of-function (expression)
MMRd: mismatch repair deficient
MSI: microsatellite instability
OS: overall survival
OTTA: ovarian tumor tissue analysis consortium
PFS: progression-free survival
TIL: tumor infiltrating lymphocytes
TMA: tissue microarray

## Acknowledgement

We thank all the study participants who contributed to this study and all the researchers, clinicians, technical and administrative staff who have made this work possible. This research was funded in part by the Janet D. Cottrelle Foundation and the Canadian Institutes of Health Research (Early Career Investigator Grant to M.S. Anglesio). K. Heinze is funded through a research scholarship by the Deutsche Forschungsgesellschaft (HE 8699/1-1). A. Talhouk is funded through a Michael Smith Foundation for Health Research Scholar Award. M. Köbel received support through the Calgary Laboratory Services research support fund (RS19-612). We thank Shuhong Liu and Young Ou (Anatomical Pathology Research Laboratory) for performing immunohistochemistry. M.S. Anglesio is funded through a Michael Smith Foundation for Health Research Scholar Award and the Janet D. Cottrelle Foundation Scholars program managed by the BC Cancer Foundation. BC’s Gynecological Cancer Research team (OVCARE) receives support through the BC Cancer Foundation and The VGH+UBC Hospital Foundation. This study uses resources provided by the Canadian Ovarian Cancer Research Consortium’s COEUR biobank funded by the Terry Fox Research Institute and managed and supervised by the Centre hospitalier de l’Université de Montréal (CRCHUM). The Consortium acknowledges contributions to its COEUR biobank from Institutions across Canada (for a full list see http://www.tfri.ca/en/research/translational-research/coeur/coeur_biobanks.aspx).

Ovarian tumor tissue analysis consortium studies were supported through independent mechanisms. The WMH study was supported by the Westmead Hospital Department of Gynaecological Oncology. The Gynaecological Oncology Biobank at Westmead, a member of the Australasian Biospecimen Network-Oncology group, was funded by the National Health and Medical Research Council Enabling Grants ID 310670 & ID 628903 and the Cancer Institute NSW Grants ID 12/RIG/1-17 & 15/RIG/1-16. The Westmead GynBiobank acknowledges financial support from the Sydney West Translational Cancer Research Centre. The Sydney West Translational Cancer Research Centre is funded by the Cancer Institute NSW. The BGS is funded by Breast Cancer Now and the Institute of Cancer Research (ICR). ICR acknowledges NHS funding to the NIHR Biomedical Research Centre. We thank the study staff, study participants, doctors, nurses, health care providers and health information sources who have contributed to the study. The SWE study, KS, CM, AL, and the GynCancer Biobank in Western Sweden is financed by Swedish Cancer foundation (KS), Swedish state under the agreement between the Swedish government and the county council, the ALF-agreement (KS) and Assar Gabrielsson foundation (CM, AL). The DOV study is funded by National Health Institute grants ID R01-CA168758, R01-CA112523 and R01-CA087538. The GER study was supported by the German Federal Ministry of Education and Research, Programme of Clinical Biomedical Research (01 GB 9401) and the German Cancer Research Center (DKFZ), and thanks Sabine Behrens for competent technical assistance. The HAW study was funded by National Institute of Health and National Cancer Institute Grants ID N01-CN-25403/CN, N01-CN-67001/CN, P30-CA-71789/CA and R01-CA-58598/CA. The HOP study was supported by National Institute of Health and National Cancer Institute Grants ID K07-CA80668, R01CA095023, and R01 CA126841, as well as by US Army Medical Research and Materiel Command DAMD17-02-1-0669 and NIH/National Center for Research Resources/General Clinical Research Center grant MO1-RR000056. MW is funded by the European Research Council Advanced Grant (H2020 BRCA-ERC under Grant Agreement No. 742432). The University of Cambridge has received salary support for author PDPP from the NHS in the East of England through the Clinical Academic Reserve. The SEA study (SEARCH) was supported through grants from Cancer Research UK (C490/A10119 C490/A10124 C490/A16561) and the UK National Institute for Health Research Biomedical Research Centres at the University of Cambridge.

## Author contribution

MSA and MK designed the study, MSA, MK, SJR, NM, KH identified specimens for the study, MK, SL, EC, PK processed samples, and performed experiments. MK, SL, and TMN reviewed pathology and generated/reviewed IHC data, KH, AT, DSC and SCYL provided statistical design input, KH drafted the manuscript. All authors revised the manuscript and approved submission of the final version.

All other author contributed through collection, curation and maintenance of respective consortia based, or local institution, collections of patient samples including recruitment and consenting of patients, clinical care, abstraction of clinical data, and updating of outcome and follow up data.

## List of supplementary files

Supplementary methods.docx

*Includes additional detail on immunohistochemistry methods, antibodies and scoring parameters*.

Table_S1-Study-Cohort.xlsx

*Includes details of the cohorts and contributing consortia-based studies*.

Supplementary tables.pdf

*Includes Tables S2-S10*.

Supplementary figures.pdf

*Includes Figures S1-S5*

