## Supplementary material for "Prognostic and Immunological Significance of ARID1A Status in Endometriosis-Associated Ovarian Carcinoma": Figures S1-S5

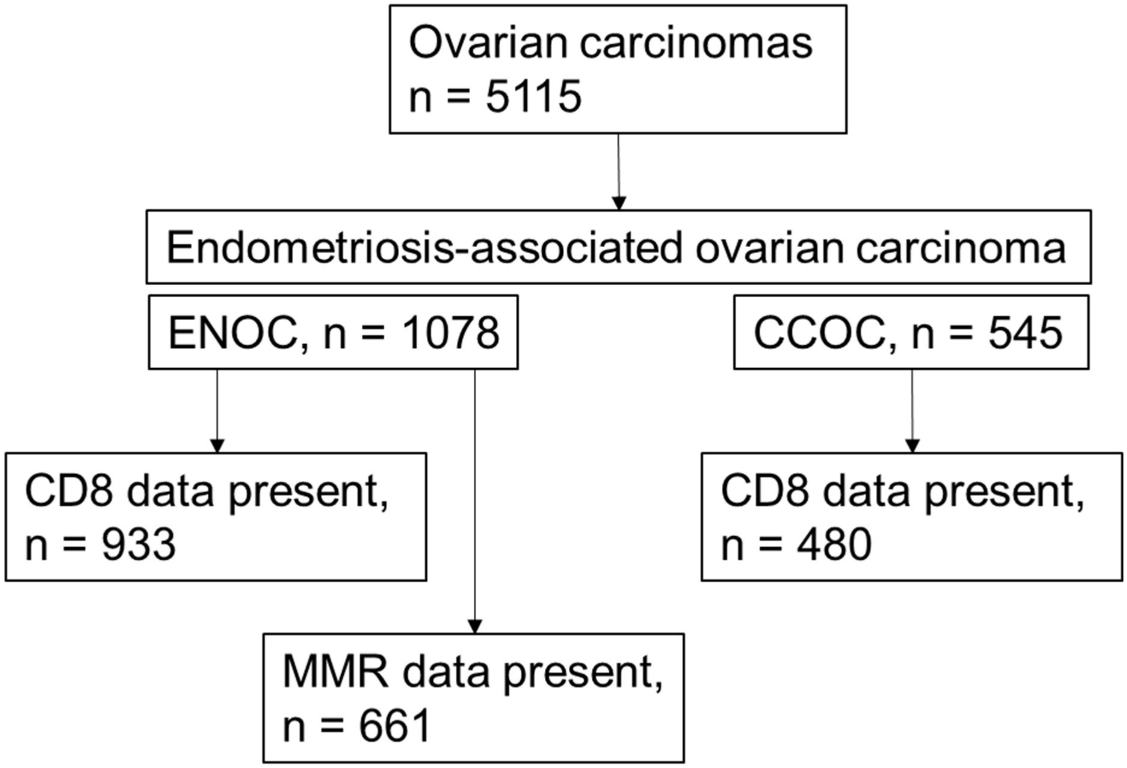


**Figure S1** Flowchart of case assignment for full and sub-cohort analysis.

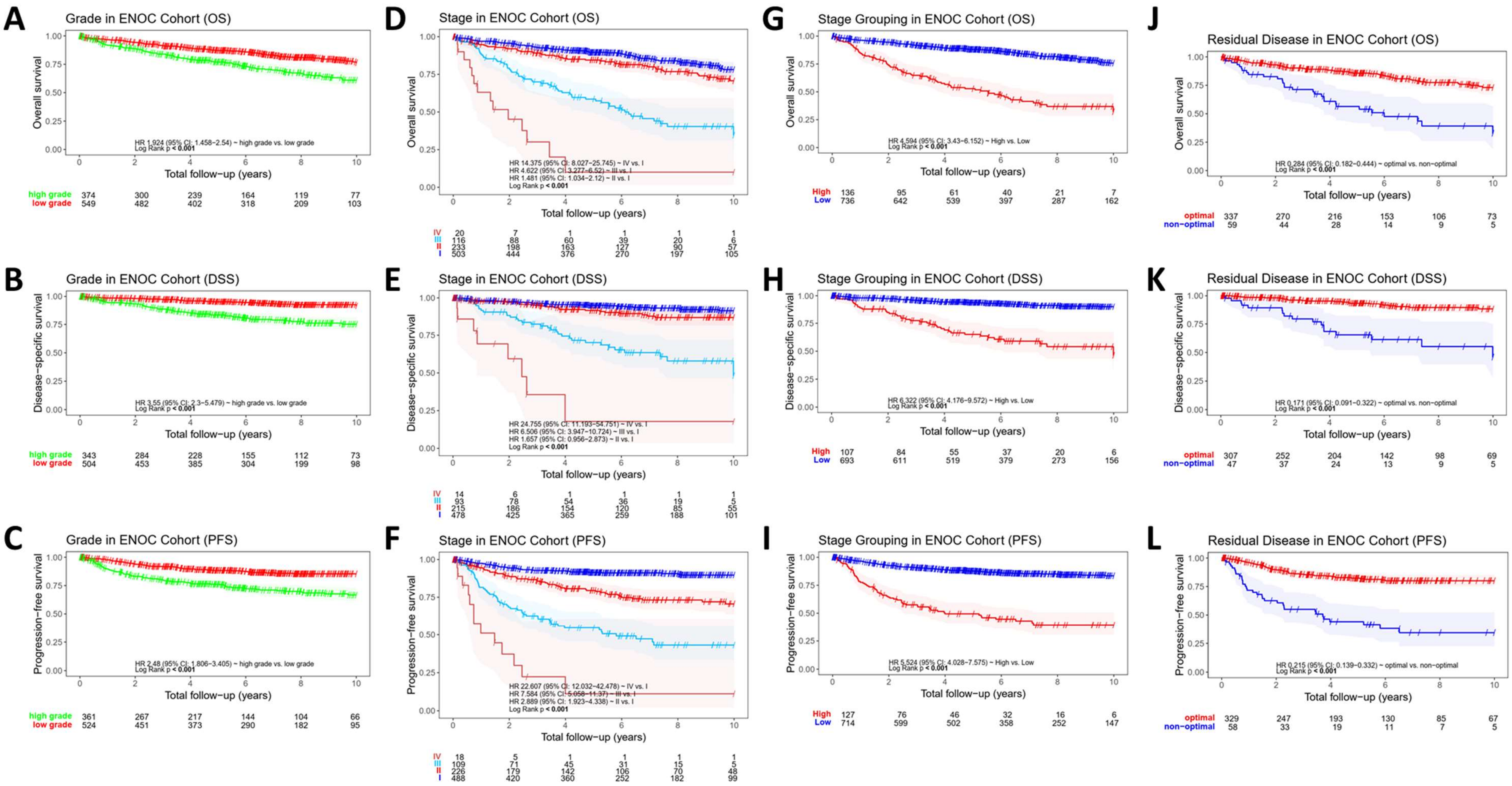


**Figure S2** Kaplan-Meier survival curves of clinicopathological features in ENOC. **A-C** Curves depicting OS (n=923), DSS (n=847) and PFS (n=611) with low vs. high grade status, **D-F** Curves depicting OS (n=872), DSS (n=800) and PFS (n=605) in respect to individual stage. **G-I** Curves depicting OS (n=872), DSS (n=800) and PFS (n=605) with low (I+II) and high (III+IV) stage. **J-L** Curves depicting OS (n=396), DSS (n=354) and PFS (n=356) in optimal and non-optimal debulked cases.

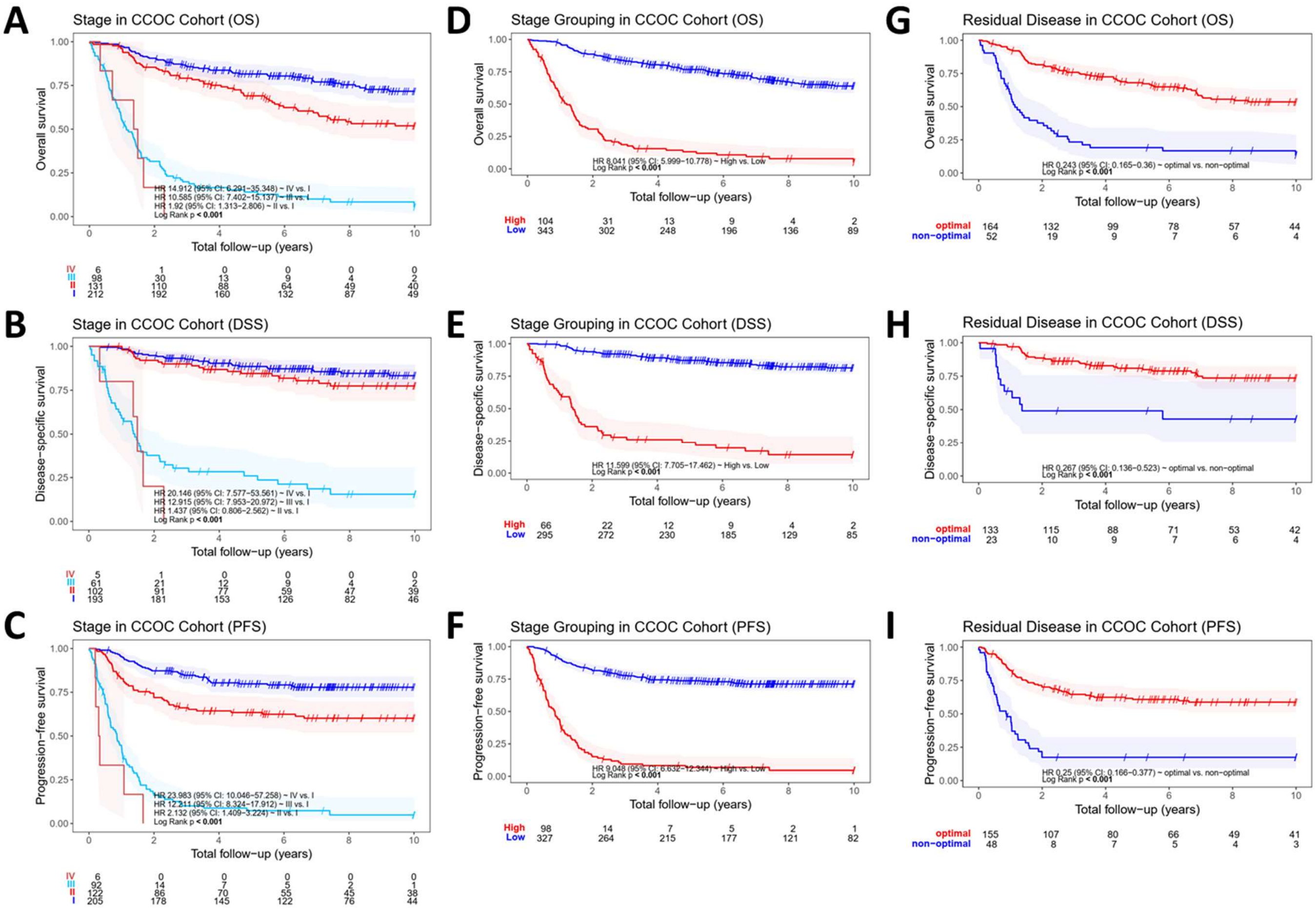


**Figure S3** Kaplan-Meier survival curves of clinicopathological features in CCOC. **A-C** Curves depicting OS (n=451), DSS (n=361) and PFS (n=313) in respect to individual stage. **D-F** Curves depicting OS (n=451), DSS (n=361) and PFS (n=313) with low (I+II) and high (III+IV) stage. **G-I** Curves depicting OS (n=216), DSS (n=156) and PFS (n=189) in optimal and non-optimal debulked cases.

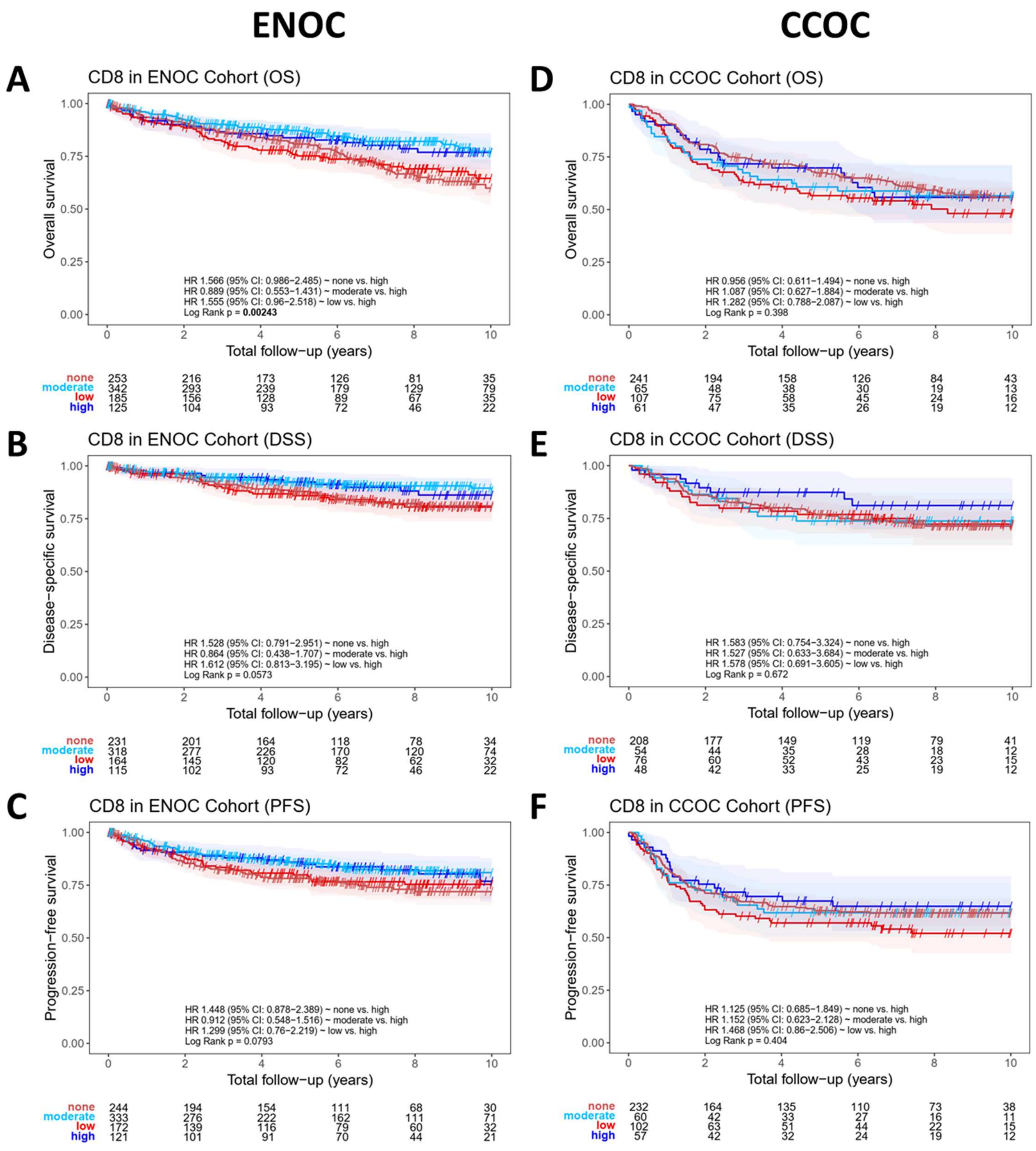


**Figure S4** Immunological influence on survival outcome in EAOC. **A-C** Kaplan-Meier survival curves depicting OS (n=905), DSS (n=828) and PFS (n=579) in respect to CD8+ TIL in ENOC. ndividual stage. **D-F** Kaplan-Meier survival curves depicting OS (n=474), DSS (n=386) and PFS (n=289) in respect to CD8+ TIL in CCOC.

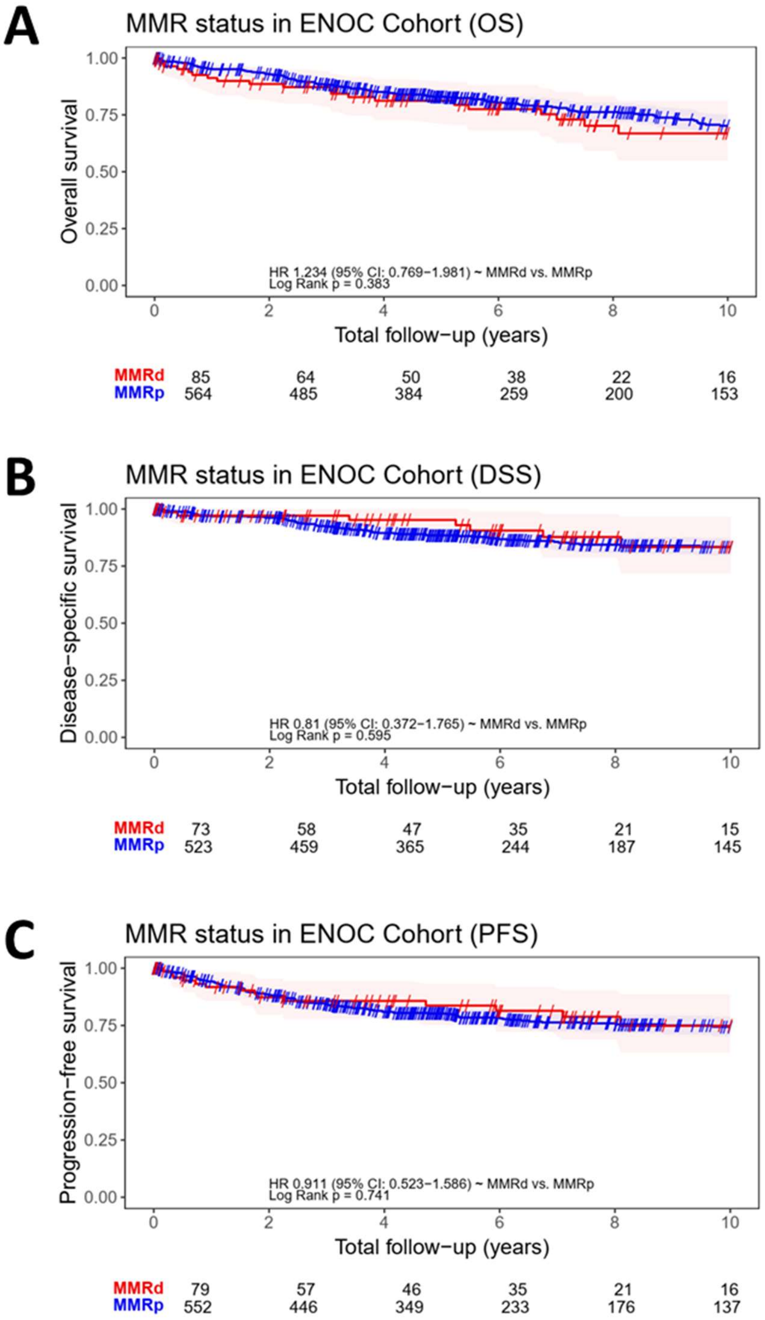


**Figure S5** MMR influence on survival outcome in ENOC. **A-C** Kaplan-Meier survival curves depicting OS (n=649), DSS (n=596) and PFS (n=554) comparing MMRd and MMRp cases.
