## Supplementary material for "Prognostic and Immunological Significance of ARID1A Status in Endometriosis-Associated Ovarian Carcinoma": Tables S2-S10

**Table S2** Cohort characteristics including all major ovarian carcinoma histological subtypes.

| Variable | Levels | AOV | BGS | CAL | DOV | GER | HAW | HOP | POC | SEA | SOC | SWE | TVA | UKO | WMH | VAN-DISC | VAN-VAL | COEUR | Heidelberg | Tübingen | Friedrichshafen | Essen | Total |
|---|---|---|---|---|---|---|---|---|---|---|---|---|---|---|---|---|---|---|---|---|---|---|---|
| **Total** | N | 550 | 105 | 112 | 1096 | 86 | 190 | 40 | 139 | 550 | 121 | 261 | 153 | 93 | 46 | 738 | 487 | 179 | 42 | 83 | 2 | 42 | 5115 |
| **Histological Subtype** | HGSC | 75 | 42 | 82 | 798 | 55 | 18 | 26 | 95 | 296 | 65 | 149 | 94 | 65 | 5 | 507 | 136 | 18 | 0 | 14 | 0 | 8 | 2548 |
| | LGSC | 2 | 0 | 24 | 13 | 0 | 0 | 0 | 6 | 11 | 2 | 19 | 5 | 4 | 0 | 16 | 8 | 1 | 0 | 0 | 0 | 0 | 111 |
| | MUC | 67 | 12 | 0 | 24 | 6 | 37 | 0 | 9 | 33 | 10 | 12 | 7 | 3 | 3 | 18 | 21 | 0 | 0 | 0 | 0 | 0 | 262 |
| | ENOC | 164 | 8 | 3 | 123 | 10 | 56 | 7 | 14 | 93 | 20 | 17 | 24 | 4 | 15 | 76 | 141 | 156 | 42 | 69 | 2 | 34 | 1078 |
| | CCOC | 152 | 1 | 0 | 76 | 1 | 35 | 1 | 0 | 64 | 3 | 12 | 17 | 10 | 14 | 77 | 82 | 0 | 0 | 0 | 0 | 0 | 545 |
| | Other/Missing | 90 | 42 | 3 | 62 | 14 | 44 | 6 | 15 | 53 | 21 | 52 | 6 | 7 | 9 | 44 | 99 | 4 | 0 | 0 | 0 | 0 | 571 |
| **Stage** | I | 226 | 5 | 4 | 258 | 15 | 38 | 12 | 13 | 0 | 32 | 95 | 38 | 25 | 27 | 130 | 185 | 94 | 24 | 43 | 2 | 14 | 1280 |
| | II | 131 | 1 | 3 | 101 | 8 | 13 | 8 | 6 | 0 | 7 | 15 | 23 | 14 | 5 | 126 | 161 | 41 | 9 | 15 | 0 | 6 | 693 |
| | III | 119 | 11 | 73 | 551 | 50 | 11 | 18 | 56 | 0 | 48 | 131 | 74 | 46 | 13 | 412 | 63 | 31 | 8 | 19 | 0 | 7 | 1741 |
| | IV | 11 | 0 | 25 | 184 | 11 | 4 | 2 | 1 | 0 | 8 | 20 | 10 | 4 | 1 | 55 | 9 | 2 | 0 | 5 | 0 | 9 | 361 |
| | Missing | 63 | 88 | 7 | 2 | 2 | 124 | 0 | 63 | 550 | 26 | 0 | 8 | 4 | 0 | 15 | 69 | 11 | 1 | 1 | 0 | 6 | 1040 |
| **Grade** | low grade | 202 | 0 | 31 | 209 | 34 | 29 | 15 | 55 | 160 | 56 | 84 | 0 | 22 | 23 | 31 | 65 | 72 | 13 | 48 | 1 | 10 | 1160 |
| | high grade | 107 | 0 | 74 | 591 | 25 | 21 | 23 | 49 | 277 | 49 | 105 | 0 | 49 | 9 | 643 | 324 | 96 | 29 | 35 | 1 | 31 | 2538 |
| | Missing | 241 | 105 | 7 | 296 | 27 | 140 | 2 | 35 | 113 | 16 | 72 | 153 | 22 | 14 | 64 | 98 | 11 | 0 | 0 | 0 | 1 | 1417 |
| **Residual Disease** | non-optimal | 103 | 0 | 84 | 0 | 0 | 12 | 0 | 0 | 0 | 0 | 76 | 54 | 0 | 10 | 20 | 4 | 24 | 2 | 17 | 0 | 2 | 408 |
| | optimal | 235 | 0 | 15 | 0 | 0 | 51 | 0 | 0 | 0 | 0 | 168 | 48 | 0 | 34 | 56 | 41 | 73 | 33 | 65 | 2 | 40 | 861 |
| | Missing | 212 | 105 | 13 | 1096 | 86 | 127 | 40 | 139 | 550 | 121 | 17 | 51 | 93 | 2 | 662 | 442 | 82 | 7 | 1 | 0 | 0 | 3846 |

**Table S3** Summary of univariable associations between ARID1A status and survival variables (OS, DSS, PFS) in ENOC (**A**) and CCOC (**B**).

**A**

| Variable | Levels | ENOC Total | ENOC PValue | ENOC Low Stage Total | ENOC Low Stage PValue | ENOC High Stage Total | ENOC High Stage PValue |
|---|---|---|---|---|---|---|---|
| **Total** | N (%) | 1078 (100%) | | 747 (100%) | | 139 (100%) | |
| **OS Status** | Censored | 745 (71%) | 0.314 | 577 (78%) | 0.808 | 59 (43%) | 0.199 |
| | Event | 299 (29%) | | 159 (22%) | | 77 (57%) | |
| | Missing | 34 | | 11 | | 3 | |
| **DSS Status** | Censored | 820 (88%) | 0.077 | 636 (92%) | 0.309 | 66 (62%) | 0.108 |
| | Event | 116 (12%) | | 57 (8%) | | 41 (38%) | |
| | Missing | 142 | | 54 | | 32 | |
| **PFS Status** | Censored | 791 (81%) | 0.511 | 614 (86%) | 0.920 | 59 (46%) | 0.503 |
| | Event | 190 (19%) | | 100 (14%) | | 68 (54%) | |
| | Missing | 97 | | 33 | | 12 | |

**B**

| Variable | Levels | CCOC Total | CCOC PValue | CCOC Low Stage Total | CCOC Low Stage PValue | CCOC High Stage Total | CCOC High Stage PValue |
|---|---|---|---|---|---|---|---|
| **Total** | N (%) | 545 (100%) | | 343 (100%) | | 105 (100%) | |
| **OS Status** | Censored | 287 (53%) | **0.048** | 217 (63%) | 0.245 | 10 (10%) | >0.999 |
| | Event | 251 (47%) | | 126 (37%) | | 94 (90%) | |
| | Missing | 7 | | 0 | | 1 | |
| **DSS Status** | Censored | 327 (75%) | 0.160 | 248 (84%) | >0.999 | 15 (23%) | 0.433 |
| | Event | 110 (25%) | | 47 (16%) | | 51 (77%) | |
| | Missing | 108 | | 48 | | 39 | |
| **PFS Status** | Censored | 309 (61%) | 0.337 | 236 (72%) | 0.501 | 9 (9%) | 0.348 |
| | Event | 194 (39%) | | 91 (28%) | | 89 (91%) | |
| | Missing | 42 | | 16 | | 7 | |

**Table S4A** Univariable association between clinicopathological variable and ARID1A status in ENOC.

**A**

| Variable | Levels | absent/subclonal | present | Total | PValue |
|---|---|---|---|---|---|
| **Total** | N (%) | 274 (25%) | 804 (75%) | 1078 (100%) | |
| **Age at Diagnosis** | Mean (sd) | 55 (11) | 57 (13) | 56 (12) | **0.012** |
| | Median (IQR) | 53 (47 - 62) | 56 (47 - 65) | 55 (47 - 64) | |
| | Missing | 0 | 3 | 3 | |
| **Stage** | I | 122 (53%) | 390 (59%) | 512 (58%) | 0.140 |
| | II | 74 (32%) | 161 (25%) | 235 (27%) | |
| | III | 29 (13%) | 90 (14%) | 119 (13%) | |
| | IV | 4 (2%) | 16 (2%) | 20 (2%) | |
| | Missing | 45 | 147 | 192 | |
| **Grade** | low grade | 139 (56%) | 426 (61%) | 565 (59%) | 0.197 |
| | high grade | 110 (44%) | 275 (39%) | 385 (41%) | |
| | Missing | 25 | 103 | 128 | |
| **Residual Disease** | non-optimal | 12 (12%) | 47 (16%) | 59 (15%) | 0.507 |
| | optimal | 86 (88%) | 253 (84%) | 339 (85%) | |
| | Missing | 176 | 504 | 680 | |

**Table S4B** Univariable association between clinicopathological variable and ARID1A status in CCOC.

**B**

| Variable | Levels | absent/subclonal | present | Total | PValue |
|---|---|---|---|---|---|
| **Total** | N (%) | 231 (42%) | 314 (58%) | 545 (100%) | |
| **Age at Diagnosis** | Mean (sd) | 55 (11) | 56 (12) | 56 (11) | 0.291 |
| | Median (IQR) | 55 (47 - 61) | 55 (48 - 65) | 55 (48 - 63) | |
| **Stage** | I | 90 (49%) | 122 (46%) | 212 (47%) | 0.712 |
| | II | 56 (30%) | 75 (28%) | 131 (29%) | |
| | III | 36 (20%) | 63 (24%) | 99 (22%) | |
| | IV | 2 (1%) | 4 (2%) | 6 (1%) | |
| | Missing | 47 | 50 | 97 | |
| **Residual Disease** | non-optimal | 20 (22%) | 32 (25%) | 52 (24%) | 0.706 |
| | optimal | 70 (78%) | 94 (75%) | 164 (76%) | |
| | Missing | 141 | 188 | 329 | |

**Table S5** Summary of univariable survival between ARID1A and CD8+ TIL status in ENOC and CCOC.

| CD8+ TIL Reference group: None | | | | | | | |
|---|---|---|---|---|---|---|---|
| | | ENOC | | | CCOC | | |
| Outcomes | Levels | Events / n | HR (95% CI) | LRT PValue | Events / n | HR (95% CI) | LRT PValue |
| OS | low | 207 / 901 | 1.06 (0.74-1.52) | **0.034** | 198 / 473 | 1.37 (0.97-1.93) | 0.377 |
| | moderate | | 0.65 (0.46-0.92) | | | 1.17 (0.76-1.8) | |
| | high | | 0.77 (0.48-1.24) | | | 1.08 (0.69-1.7) | |
| DSS | low | 99 / 824 | 1.1 (0.65-1.86) | 0.198 | 95 / 385 | 1 (0.59-1.7) | 0.522 |
| | moderate | | 0.65 (0.38-1.09) | | | 0.97 (0.53-1.79) | |
| | high | | 0.75 (0.38-1.48) | | | 0.59 (0.28-1.25) | |
| PFS | low | 165 / 865 | 0.88 (0.58-1.34) | 0.086 | 174 / 450 | 1.35 (0.94-1.93) | 0.324 |
| | moderate | | 0.63 (0.43-0.93) | | | 1.01 (0.63-1.62) | |
| | high | | 0.66 (0.39-1.1) | | | 0.87 (0.53-1.44) | |

**Table S6** Summary of univariable association of MMR protein status and ARID1A loss-of function in ENOC.

| Variable | ENOC | | | Low stage ENOC | | |
|---|---|---|---|---|---|---|
| N (%) | ARID1A loss | ARID1A present | pValue | ARID1A loss | ARID1A present | pValue |
| MLH1 | 18 (11%) | 25 (5%) | **0.015** | 14 (10%) | 16 (4%) | **0.013** |
| MSH2 | 8 (5%) | 14 (3%) | 0.307 | 5 (4%) | 10 (3%) | 0.673 |
| MSH6 | 14 (9%) | 23 (5%) | 0.081 | 9 (7%) | 17 (4%) | 0.349 |
| PMS2 | 19 (12%) | 16 (3%) | **<0.001** | 14 (11%) | 10 (3%) | **<0.001** |
| MMRd | 36 (22%) | 51 (10%) | **<0.001** | 26 (19%) | 35 (9%) | **0.003** |
| MMRp | 131 (78%) | 443 (90%) | | 112 (81%) | 360 (91%) | |
| MMR missing | 107 | 310 | | 58 | 156 | |

**Table S7** Univariable associations between MMR status and survival variables (OS, DSS, PFS) in ENOC.

| Variable | Levels | MMRp | MMRd | Total | PValue |
|---|---|---|---|---|---|
| **Total** | N (%) | 574 (53%) | 87 (8%) | 1078 (100%) | |
| **OS Status** | Censored | 415 (74%) | 62 (73%) | 477 (73%) | >0.999 |
| | Event | 149 (26%) | 23 (27%) | 172 (27%) | |
| | Missing | 10 | 2 | 12 | |
| **DSS Status** | Censored | 455 (87%) | 65 (89%) | 520 (87%) | 0.762 |
| | Event | 68 (13%) | 8 (11%) | 76 (13%) | |
| | Missing | 51 | 14 | 65 | |
| **PFS Status** | Censored | 434 (79%) | 63 (80%) | 497 (79%) | 0.935 |
| | Event | 118 (21%) | 16 (20%) | 134 (21%) | |
| | Missing | 22 | 8 | 30 | |

**Table S8** Relationship between ARID1A mutation type and MMR status, based on [10,61,62]. A 2-tailed Fisher test showed significant correlation between indel ARID1A LOF mutation and MSI and MMRd status.

| ARID1A(MUTANT/LOF) | Indel | Non-INDEL(SNV, Splice, fus, CNC) |
|---|---|---|
| MSI/MMRd | 5 | 1 |
| MSS/MMRp | 1 | 8 |

Fisher (2-tail) test p=0.0110

**Table S9** Univariable analysis in ENOC MMRd cohort. **A** Univariable associations between ARID1A status and clinicopathological variables. **B** Univariable survival between ARID1A status and clinicopathological and molecular variables.

**A**

| Variable | Levels | absent/subclonal | present | Total | PValue |
|---|---|---|---|---|---|
| **Total** | N (%) | 36 (41%) | 51 (59%) | 87 (100%) | |
| **Age at Diagnosis** | Mean (sd) | 52 (11) | 56 (13) | 54 (12) | 0.129 |
| | Median (IQR) | 51 (43 - 54) | 53 (46 - 61) | 52 (45 - 59) | |
| **Grade** | low grade | 15 (42%) | 23 (47%) | 38 (45%) | 0.793 |
| | high grade | 21 (58%) | 26 (53%) | 47 (55%) | |
| | Missing | 0 | 2 | 2 | |
| **Stage** | I | 16 (46%) | 21 (47%) | 37 (46%) | 0.876 |
| | II | 10 (29%) | 14 (31%) | 24 (30%) | |
| | III | 7 (20%) | 9 (20%) | 16 (20%) | |
| | IV | 2 (6%) | 1 (2%) | 3 (4%) | |
| | Missing | 1 | 6 | 7 | |

**B**

| | # of events / n | | Hazard Ratio (95% CI) | LRT P-value |
|---|---|---|---|---|
| **Age (reference group:NA)** | | | | |
| OS | 20 / 85 | | 1.16 (0.93-1.45) | 0.164 |
| DSS | 7 / 73 | | 1.05 (0.78-1.4) | 0.749 |
| PFS | 14 / 79 | | 1.15 (0.91-1.47) | 0.218 |
| **Stage (reference group:I)** | | | | |
| OS | 16 / 79 | II | 0.53 (0.1-2.73) | **0.010** |
| | | III | 0.76 (0.15-3.83) | |
| | | IV | 1.64e+09 (0-Inf) | |
| DSS | 7 / 71 | II | 1.58 (0.23-10.92) | 0.139 |
| | | III | 1.09 (0.11-11.24) | |
| | | IV | 4.1e+09 (0-Inf) | |
| PFS | 12 / 75 | II | 0.72 (0.13-4.12) | **0.007** |
| | | III | 2.58 (0.51-13.02) | |
| | | IV | 3.9e+09 (0-Inf) | |
| **Grade (reference group:low grade)** | | | | |
| OS | 19 / 84 | high grade | 0.42 (0.14-1.27) | 0.110 |
| DSS | 7 / 73 | high grade | 0.7 (0.15-3.21) | 0.649 |
| PFS | 13 / 78 | high grade | 0.79 (0.24-2.6) | 0.698 |
| **Residual Disease (reference group:non-optimal)** | | | | |
| OS | 12 / 47 | optimal | 0.41 (0.07-2.46) | 0.357 |
| DSS | 3 / 38 | optimal | 0 (0-Inf) | **0.041** |
| PFS | 8 / 43 | optimal | 0.28 (0.04-2.22) | 0.242 |
| **ARID1A (reference group:absent/subclonal)** | | | | |
| OS | 20 / 85 | present | 0.81 (0.3-2.16) | 0.671 |
| DSS | 7 / 73 | present | 0.77 (0.14-4.23) | 0.766 |
| PFS | 14 / 79 | present | 1.14 (0.35-3.72) | 0.822 |
| **CD8+ (reference group:none)** | | | | |
| OS | 20 / 83 | low | 1.49 (0.25-8.81) | 0.924 |
| | | moderate | 1.03 (0.18-5.86) | |
| | | high | 1.54 (0.26-9.08) | |
| DSS | 7 / 71 | low | 0.46 (0.03-7.85) | 0.792 |
| | | moderate | 1.28 (0.13-13.11) | |
| | | high | 1.31 (0.11-16.13) | |
| PFS | 14 / 77 | low | 1.18 (0.12-11.78) | 0.984 |
| | | moderate | 1.45 (0.16-13.29) | |
| | | high | 1.37 (0.14-13.36) | |

**Table S10** Univariable survival between ARID1A status and clinicopathological and molecular variables in ENOC MMRp cohort.

| | # of events / n | | Hazard Ratio (95% CI) | LRT P-value |
|---|---|---|---|---|
| **Age (reference group:NA)** | | | | |
| OS | 121 / 563 | | 1.07 (1-1.15) | 0.051 |
| DSS | 66 / 522 | | 1.15 (1.04-1.28) | **0.005** |
| PFS | 115 / 551 | | 1.27 (1.17-1.39) | **< 0.001** |
| **Stage (reference group:I)** | | | | |
| OS | 113 / 535 | II | 1.63 (1.01-2.63) | **< 0.001** |
| | | III | 8.23 (5.1-13.28) | |
| | | IV | 9.49 (3.83-23.52) | |
| DSS | 59 / 497 | II | 2.02 (1.02-4.03) | **< 0.001** |
| | | III | 12.25 (6.26-23.99) | |
| | | IV | 15.5 (4.3-55.9) | |
| PFS | 106 / 524 | II | 3.8 (2.27-6.35) | **< 0.001** |
| | | III | 12.5 (7.28-21.47) | |
| | | IV | 19.27 (7.28-51.02) | |
| **Grade (reference group:low grade)** | | | | |
| OS | 109 / 536 | high grade | 2.45 (1.62-3.69) | **< 0.001** |
| DSS | 58 / 499 | high grade | 5.32 (2.67-10.6) | **< 0.001** |
| PFS | 103 / 525 | high grade | 2.28 (1.5-3.46) | **< 0.001** |
| **Residual Disease (reference group:non-optimal)** | | | | |
| OS | 63 / 276 | optimal | 0.15 (0.09-0.27) | **< 0.001** |
| DSS | 33 / 250 | optimal | 0.11 (0.05-0.24) | **< 0.001** |
| PFS | 64 / 271 | optimal | 0.12 (0.07-0.2) | **< 0.001** |
| **ARID1A (reference group:absent/subclonal)** | | | | |
| OS | 121 / 563 | present | 1.16 (0.73-1.87) | 0.521 |
| DSS | 66 / 522 | present | 1.52 (0.77-3.02) | 0.209 |
| PFS | 115 / 551 | present | 1.19 (0.75-1.9) | 0.450 |
| **CD8+ (reference group:none)** | | | | |
| OS | 112 / 541 | low | 1.09 (0.66-1.81) | 0.095 |
| | | moderate | 0.62 (0.38-0.99) | |
| | | high | 0.74 (0.36-1.51) | |
| DSS | 61 / 502 | low | 1.1 (0.57-2.1) | **0.038** |
| | | moderate | 0.47 (0.24-0.93) | |
| | | high | 0.47 (0.17-1.32) | |
| PFS | 109 / 530 | low | 0.84 (0.5-1.42) | 0.095 |
| | | moderate | 0.57 (0.35-0.91) | |
| | | high | 0.61 (0.31-1.2) | |
