## Supplementary methods for "Prognostic and Immunological Significance of ARID1A Status in Endometriosis-Associated Ovarian Carcinoma"

**Supplementary method**

*ARID1A staining and scoring*

ARID1A staining was performed in two assay setup variations in 2016 and 2018. The first IHC protocol was done with rabbit polyclonal antibody from Sigma (cat# HPA005456) at 1:150 or 1:200 dilution with 10-X-10 incubation. The second protocol was performed with rabbit monoclonal antibody clone EPR13501 from Abcam (cat# ab182560) and a 1:2000 dilution. In both protocol the staining was completed using the Dako Omnis platform, a pretreatment with TRS high and detection HRP based using 3,3’-diaminobenzidine (DAB).

ARID1A scoring was done by MK (main) and SL (TVA for interobserver reproducibility) using the following scoring system:

| 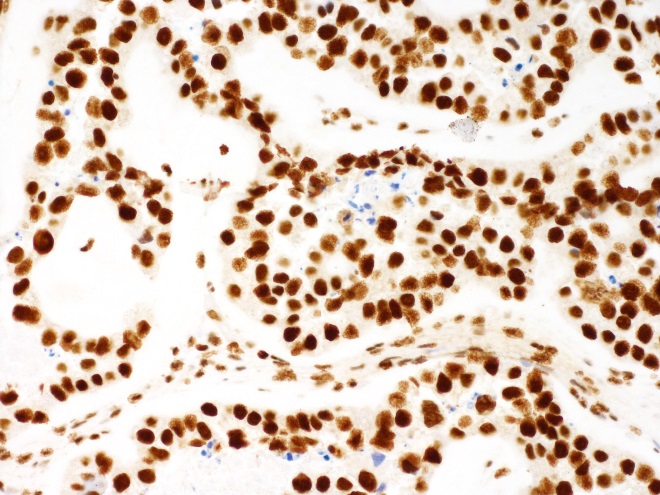 | 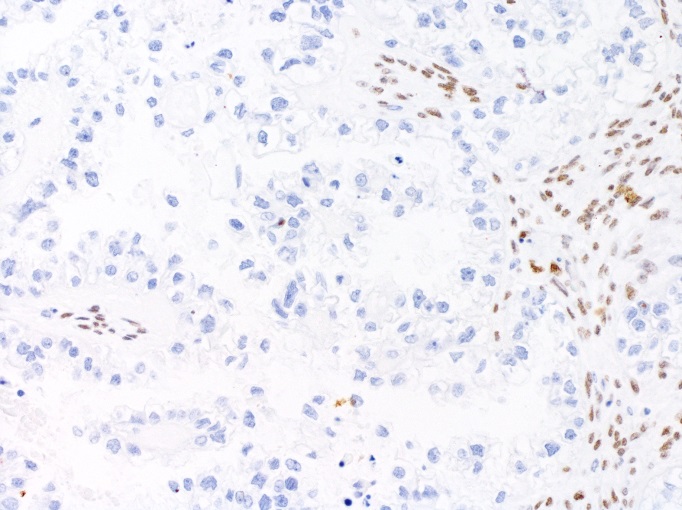 |
| --- | --- |
| Score 1 = present | Score 0 = absent |
| 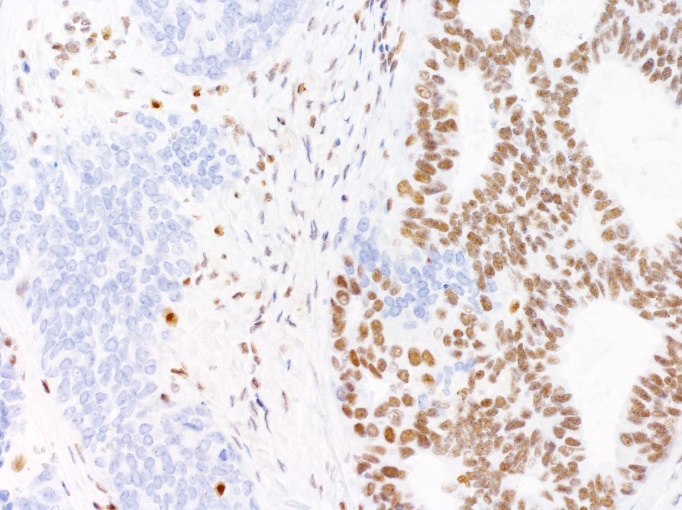 | 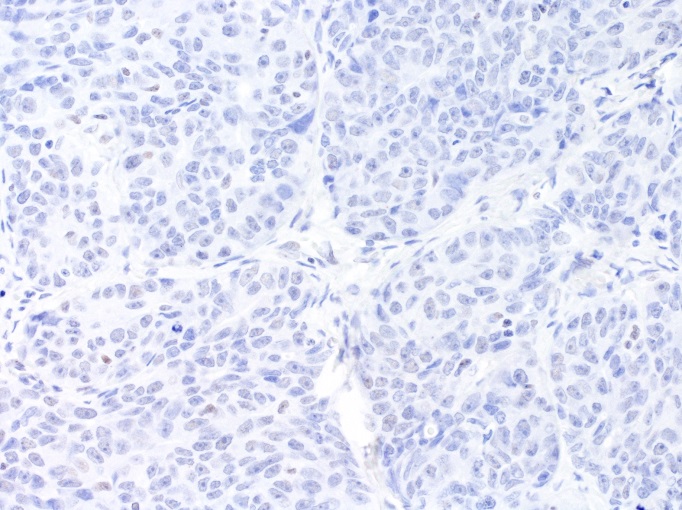 |
| Score 2 = Subclonal loss | Score 8 = no staining and lack of internal control |

*MMR staining*

MMR protein staining was achieved using the Dako Omnis platform with ready-to-use clone ready-to-use clone ES05 (Dako) for MLH1; clone EP51 (Dako) for PMS2; clone FE11 (Dako) for MSH2 and clone EP49 (or EPR3945; Dako) for MSH6 [55,60]. Dependent on retained nuclear staining in non-malignant cells as internal control, cases were considered MMR deficient in absence of nuclear staining in malignant cells.
